# Heterozygous deleterious MUTYH variants as a driver for tumorigenesis

**DOI:** 10.1101/2021.06.09.21258588

**Authors:** Rodrigo Araujo Sequeira Barreiro, Jorge Sabbaga, Benedito M. Rossi, Maria Isabel W Achatz, Anamaria A Camargo, Paula F Asprino, Pedro A F Galante

## Abstract

MUTYH is a glycosylase involved in the base excision repair of the DNA. Biallelic mutations in the MUTYH gene cause the autosomal recessive condition known as MUTYH-associated adenomatous polyposis and increase colorectal cancer risk. However, the cancer risk associated with germline variants in individuals carrying only one MUTYH defective allele is controversial and based on studies involving few samples. Here, we described a comprehensive investigation of monoallelic deleterious MUTYH carriers among approximately 10,400 patients across 33 different tumor types and more than 117 thousand samples of normal individuals. Our results indicate MUTYH deficiency in heterozygosity can lead to tumorigenesis through a mechanism of Loss Of Heterozygosity (LOH) of the functional MUTYH allele. We confirmed that the frequency of damaging MUTYH monoallelic variant carriers is higher in individuals with cancer than in the general population, though its frequency is not homogeneous among tumor types. We also demonstrate that MUTYH related mutational signature is elevated only in those patients with loss of the functional allele. We also find that MUTYH characteristic base substitution (C>A) increases stop codon generation and we identify key genes affected during tumorigenesis. In conclusion, we propose that deleterious germline monoallelic MUTYH variant carriers are at a higher risk of developing tumors, especially those types with frequent LOH events, such as adrenal adenocarcinoma.

## Background

Inefficient DNA repair is a critical driving force behind cancer establishment, progression and evolution. MUTYH, a gene located at chromosome locus 1p34.3-p32.1, encodes a glycosylase involved in base excision repair (BER) [1]. DNA glycosylases comprise a family of conserved enzymes known to recognize DNA lesions, cleaving the bond between the deoxyribose sugar and a mismatched or modified base. Each DNA glycosylase executes specific roles, but there are overlapping specificities among them [2].

The MUTYH’s glycosylase removes adenines misincorporated opposite to 8-oxoG, resulting from erroneous replication of DNA containing 8-oxoG that has escaped the action of other glycosylase, OGG1 [3]. Deficiency of the BER mechanism implies accumulation of specific somatic mutations, leaving behind a characteristic error trace [4,5]. Biallelic mutations in the *MUTYH* gene are known to cause the autosomal recessive condition known as MUTYH-associated adenomatous polyposis (MAP) [6,7]. This condition is characterized by multiple colorectal adenomas and is responsible for a small fraction (<1%) of colorectal cancers [8,9]. The frequency of MUTYH pathogenic germline variants in the general population has been estimated to be as high as 1:45, with specific founder mutations overrepresented [10,11]. Despite some evidence indicating predisposition to cancer types, such as breast, ovary and bladder, there are only a few studies associating *MUTYH* pathogenic germline variants and cancer formation [12]. Moreover, the cancer risk associated with carrying only one *MUTYH* defective variant is controversial and no molecular mechanisms linking *MUTYH* pathogenic germline variants and cancer have been proposed.

Here, we comprehensively investigated approximately 10.4 thousand samples from 33 cancer types to define the risk and the putative molecular mechanism by which carriers of monoallelic germline deleterious *MUTYH* develop cancer. We show that, although rare, *MUTYH* deficiency in heterozygosity can lead to tumorigenesis through a mechanism of Loss Of Heterozygosity (LOH) of the functional allele. This malignant transformation pathway is not restricted to a specific tumor type, but seems to be increased in cancer types frequently presenting LOH events.

## 2 RESULTS

### 2.1 MUTYH deleterious germline variants in the TCGA samples

In order to find individuals carrying germline *MUTYH* variants we analyzed germline mutation data from 10,389 patients available in the The Cancer Genome Atlas (TCGA) (Figure 1A). It is a very rich data comprehending more than 1,64 billion germline variants in several genes [13]. In *MUTYH*, we found 359 variants (Table S1). We filtered the *MUTYH* set of variants to exclude those which may not have a functional impact on the protein activity (see methods). Briefly, we selected variants that: (1) were classified as pathogenic or likely pathogenic on ClinVar[14]; (2) were a frameshift insertion or deletion InDel; (3) or had a damaging status on at least two of three prediction softwares: SIFT[15], PolyPhen[16] and REVEL[17]. After applying these filters, we gathered a total of 49 distinct *MUTYH* germline variants presenting in a monoallelic fashion on 224 TCGA’s patients (Table S2). No individual bearing two defective germline *MUTYH* variants was identified in this dataset. The variants set spanned all of MUTYH’s exons and the majority (37 out of 49) of the variants were represented by single carriers unique to one individual while only 3 variants were found on more than 20 different patients (Table S2; Figure 1B).

**Figure 1.**
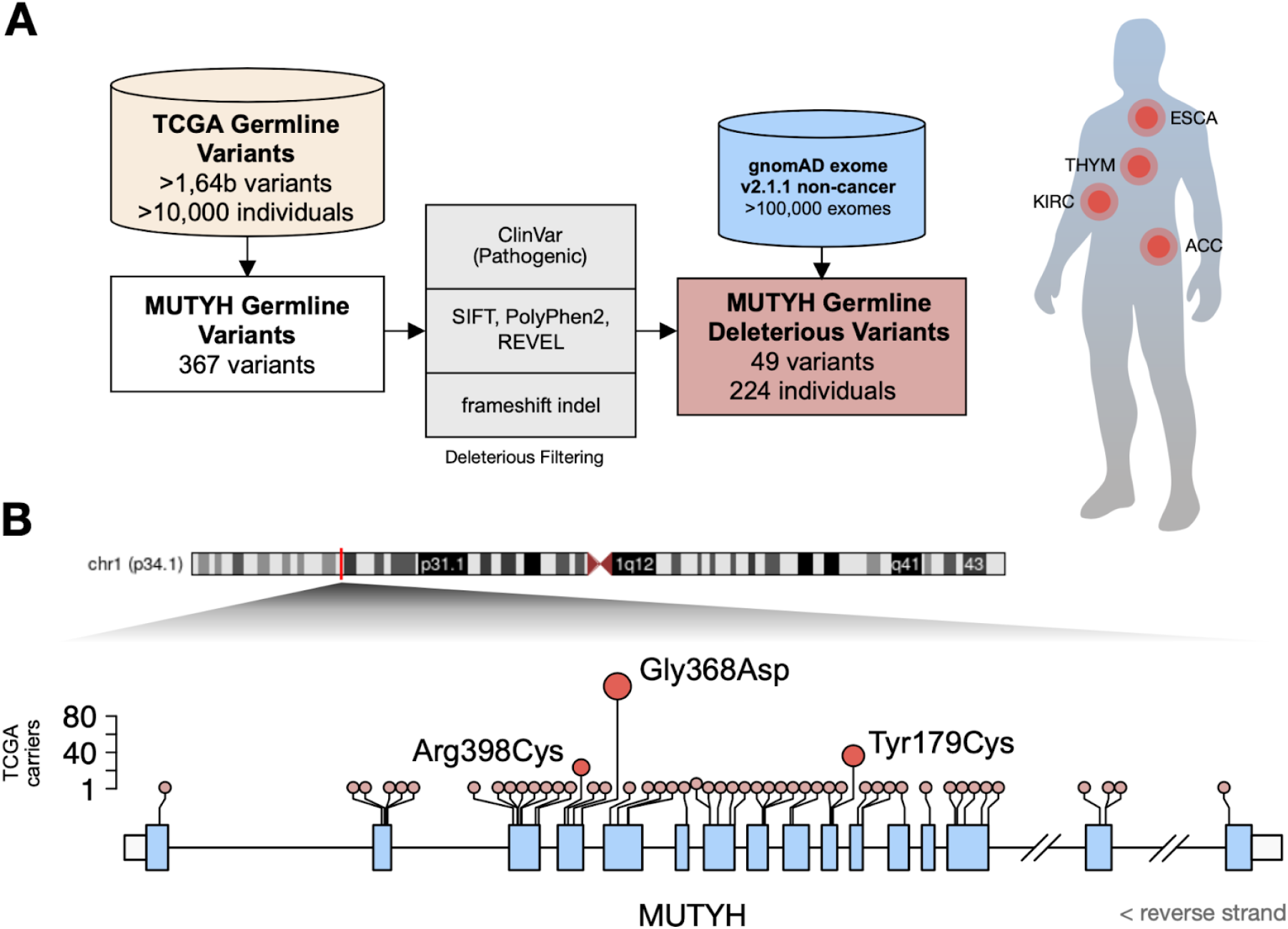
Deleterious MUTYH germline variants and their occurrence in TCGA. (A) Schematic representation of our approach to investigate MUTYH deleterious germline mutations in approximately 10 thousand cancer samples from TCGA. (B) MUTYH genomic region representation and damaging variants observed. Light blue boxes illustrate the coding exons while white boxes represent the 3’ and 5’ UTR regions. The circles symbolize the damaging mutations found by our pipeline. The size and height of the circle represent the frequency of the variant carriers in the TCGA cohort.

Our control group of non oncologic patients was based on approximately 117.7 thousand individuals from the gnomAD (v2.1.1, non-cancer subset)[18], excluding those samples from cancer databases such as the TCGA. In this cohort we found 1,695 (1.44%) monoallelic deleterious MUTYH carriers, considering the same MUTYH variant set that we extracted from the TCGA cohort (Table S3).

### 2.2 Germline deleterious MUTYH variants are enriched in cancers

Next, we investigated the frequency of the MUTYH germline deleterious variants in tumors and normal samples. Considering more than 117.7 thousands individuals from gnomAD, we found 1.43% carrying germline deleterious MUTYH variants while TCGA samples presented significantly more *MUTYH* deleterious variants, 2.1% (p-value = 0.0001; Fisher-Exact test), Figure 2A.

**Figure 2.**
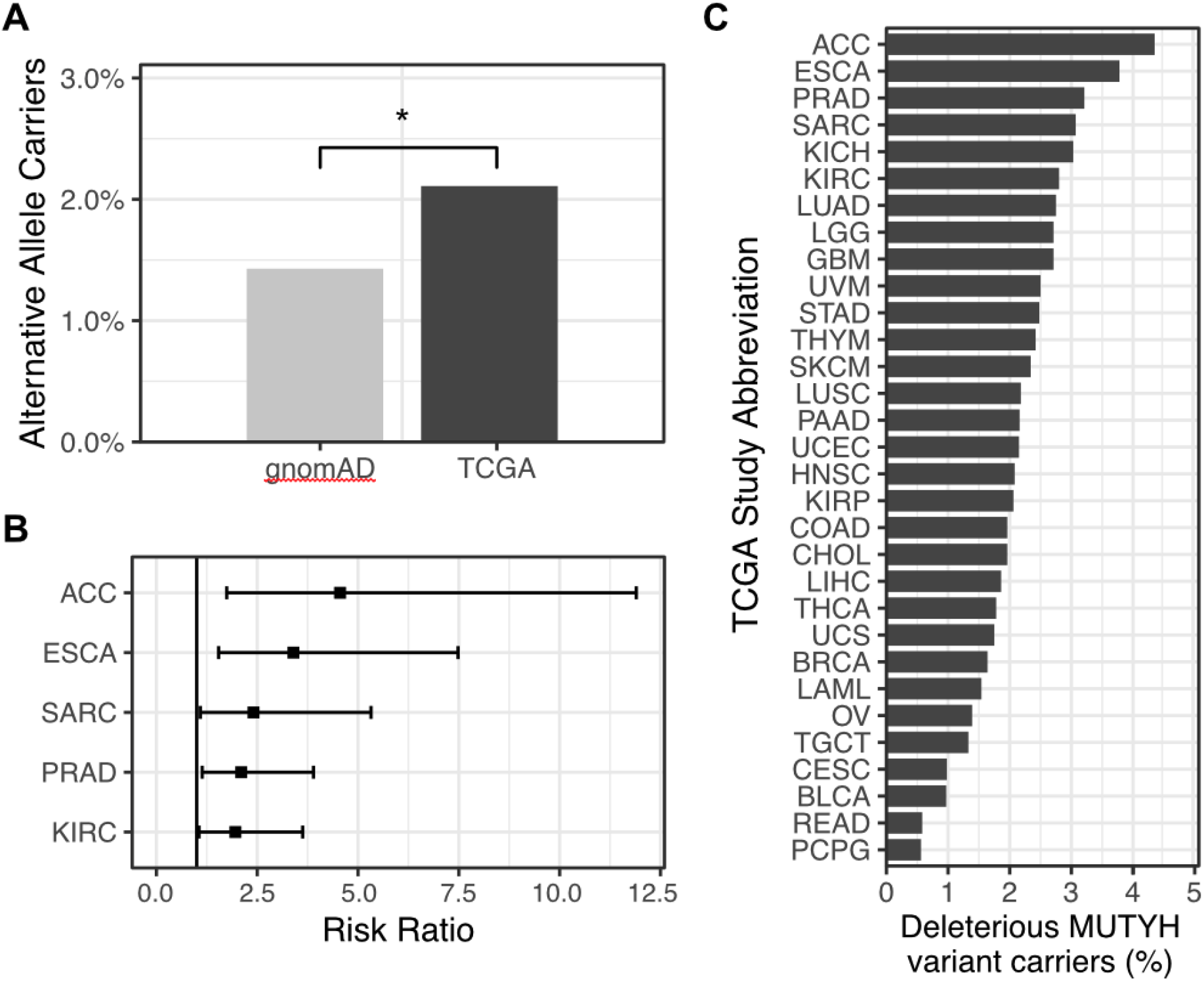
Germline deleterious MUTYH variants are enriched in some cancer cohorts. (A) Percent of monoallelic germline deleterious MUTYH variant carriers on gnomAD and TCGA cohorts. (B) Risk Ratio with 95% confidence interval for tumor types within germline deleterious MUTYH variant carriers. (C) Proportion of carriers on TCGA cohort by tumor type (abbreviations according to TCGA studies: ACC: Adrenocortical carcinoma; ESCA: Esophageal carcinoma; PRAD: Prostate adenocarcinoma; SARC: Sarcoma; KICH: Kidney chromophobe; KIRC: Kidney renal clear cell carcinoma; LUAD: Lung adenocarcinoma; LGG: Brain lower grade glioma; GBM: Glioblastoma multiforme; UVM: Uveal melanoma; STAD: Stomach adenocarcinoma; THYM: Thymoma; SKCM: Skin cutaneous melanoma; LUSC: Lung squamous cell carcinoma; PAAD; Pancreatic adenocarcinoma; UCEC: Uterine corpus endometrial carcinoma; HNSC: Head and neck squamous cell carcinoma; KIRP: Kidney renal papillary cell carcinoma; COAD: Colon adenocarcinoma; CHOL: Cholangiocarcinoma; LIHC: Liver hepatocellular carcinoma; THCA: Thyroid carcinoma; UCS: Uterine carcinosarcoma; BRCA: Breast invasive carcinoma; LAML: Acute myeloid leukemia; OV: Ovarian serous cystadenocarcinoma; TGCT: Testicular germ cell tumors; CESC: Cervical squamous cell carcinoma and endocervical adenocarcinoma; BLCA: Bladder urothelial carcinoma; READ: Rectum adenocarcinoma; PCPG: Pheochromocytoma and paraganglioma). * p-value < 0.0001.

In order to determine whether monoallelic MUTYH carriers had a higher risk of developing any type of cancer, we calculated the risk ratio comparing the TCGA and the gnomAD cohorts (Figure 2B). We found that monoallelic MUTYH carriers had a higher Risk Ratio of 4.55 (95% CI, 1.74-11.9) for developing adrenocortical carcinoma (ACC), 3.4 (95% CI: 1.54-7.48) for esophageal carcinoma (ESCA), 2.4 (95% CI: 1.09-5.32) for sarcoma (SARC), 2.1 (95% CI: 1.14-3.89) for prostate adenocarcinoma (PRAD) and 1.96 (95% CI: 1.06-3.62) for kidney renal clear cell carcinoma (KIRC), Figure 2B.

We also explored the proportion of carriers and non-carriers across all tumor types from TCGA (Figure 2C). Our data indicates that monoallelic MUTYH frequency is not homogeneous among tumor types, being most prevalent in adrenocortical carcinoma (4.35% of individuals) and in esophageal carcinoma (3.78% of individuals), while colon (1.96% of individuals) and rectum adenocarcinoma (0.58% of individuals) display modest representations. Importantly, somatic mutations in MUTYH were not considered here because they are found within 7 samples only (Table S4).

### 2.3 Loss of heterozygosity in tumors with germline deleterious variants in MUTYH

During tumorigenesis, tumor suppressor genes frequently go through loss of heterozygosity (LOH), as described by Knudson’s two hit hypothesis [19]. We investigate LOH in the MUTYH region for the alternative MUTYH variant carriers using TCGA LOH data [20]. Considering 224 deleterious MUTYH carriers, we found 27 patients (12.05%) presenting LOH in the MUTYH region, according to the ABSOLUTE pipeline [21], Figure 3A. Next, we evaluated the mutant allele frequency (MAF) of MUTYH in LOH and No-LOH TCGA carriers using TCGA’s RNA sequencing (RNA-seq) information. As expected, carriers without LOH had around 50% MAF, while for the LOH group, the MAF shifted for the low or high frequency extremities (Figure 3B). In order to investigate the patients with more confidence of loss of the MUTYH functioning copy, we grouped the samples with LOH and High MAF (MAF > 60%, 11 of the 27 patients) for the following analyses. Considering the adrenocortical carcinoma (ACC), three out of four samples with LOH and MUTYH variants presented MAF above 60%, indicating higher expression of the variant allele, while no read reporting the variant allele was found in one sample.

**Figure 3.**
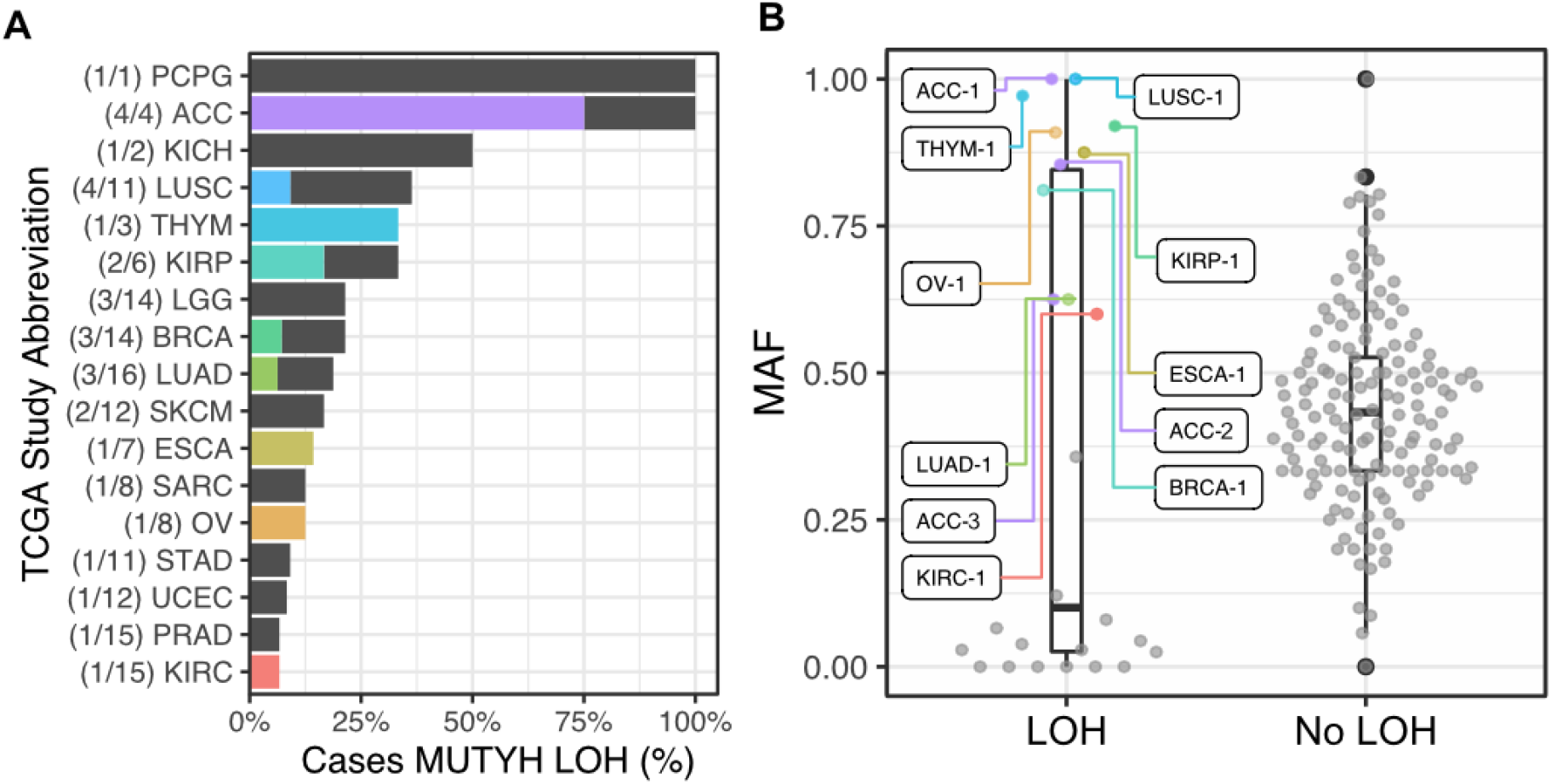
MUTYH loss of heterogeneity and gene expression. (A) Proportion of TCGA damaging MUTYH carriers with loss of heterogeneity (LOH) for MUTYH region according to ABSOLUTE analysis. In parenthesis are the number of MUTYH variant carriers with LOH followed by MUTYH variant carriers. Samples with MAF > 0.6 on the RNA-seq are highlighted. (B) Mutant Allele Frequency (MAF) from MUTYH variant carriers RNA-seq data. Highlighted patients represent carriers with LOH for MUTYH region and MAF > 0.6.

### 2.4 Tumors with MUTYH variant have specific mutational profiles

In terms of molecular function, MUTYH activity is related to the removal of misincorporated adenines opposite to 8-oxoG. Malfunctioning of MUTYH results in enhanced Cytosine to Adenine (C>A) transversions. Thereby, we sought for mutational signatures in those tumors carrying deleterious MUTYH variants. First, we found the contribution of mutational signature 18, from Mutational Signature v2 [22]. This signature, characteristic of MUTYH deficiency [23,24], was over represented only in the set of MUTYH monoallelic carriers with LOH and high MAF, while monoallelic carriers alone had similar number to the non-carriers in TCGA’s cohort (Figure 4A). The number of patients with any contribution of this signature was elevated only in the MUTYH variant carriers with LOH and high MAF (90.9%, 10 out of 11 patients). Next, we evaluated the C>A substitutions in all neighboring nucleotides contexts (Figure 4B). In general, C>A mutations were more prevalent in MUTYH variant carriers, yet T(C>A)T and G(C>A)A were the most relevant differences. At last, we gathered the seven most prevalent signatures in the MUTYH variant carriers with LOH and high MAF (Figure 4C). We observed another C>A signature also present in this set, signature 29, while signature 04 had also contributed to some of the mutational profiles.

**Figure 4.**
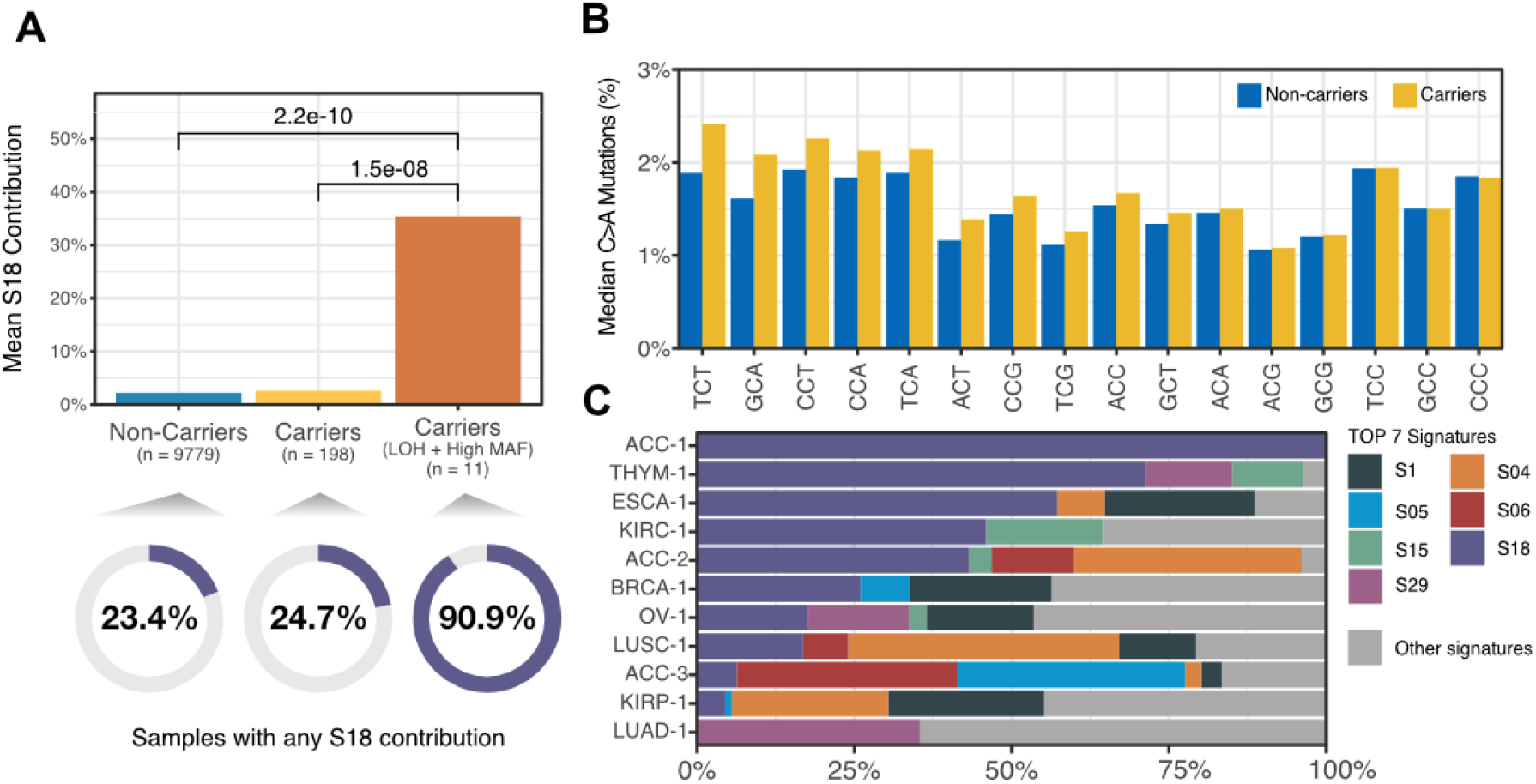
Impact of MUTYH germline variants in tumor mutational profile of TCGA cohort. (A) Contribution of COSMIC Mutational Signature 18 in non-carriers, MUTYH variant carriers and carriers with LOH and High MAF (RNA-seq MAF > 0.6). In the bottom, the proportion of samples with any Signature 18 contribution in the patient’s somatic mutational set. (B) Proportion of C>A mutations with genomic context for carriers (yellow) and non-carriers (blue). (C) COSMIC Mutational Signature composition por the 11 damaging MUTYH carriers with LOH and High MAF.

### 2.5 Stop gain generation as a potential tumorigenic process on MUTYH carriers

As previously exposed, C>A transversions are favored in tumors from MUTYH carriers that have undergone loss of the functional allele. As Adenine is the most prevalent nitrogenous base in stop codons composition (TAA, TGA and TAG), we investigate the stop generation within the TCGA cohort for non-MUTYH variant carriers, carriers and carriers with LOH and high MAF. First we evaluated the probability of all possible mutations to generate stop gain codons by both theoretical and simulation approaches (Figure S1). As expected C>A along with T>A were the two mutations with most stop gain generation potential. Next, we analyzed the stop-gain proportion in the TCGA cohort. As occurred with the signature analysis, an elevated proportion of stop gain mutations is observed only in the MUTYH carriers with the loss of the wild-type copy, Figure 5A.

**Figure 5.**
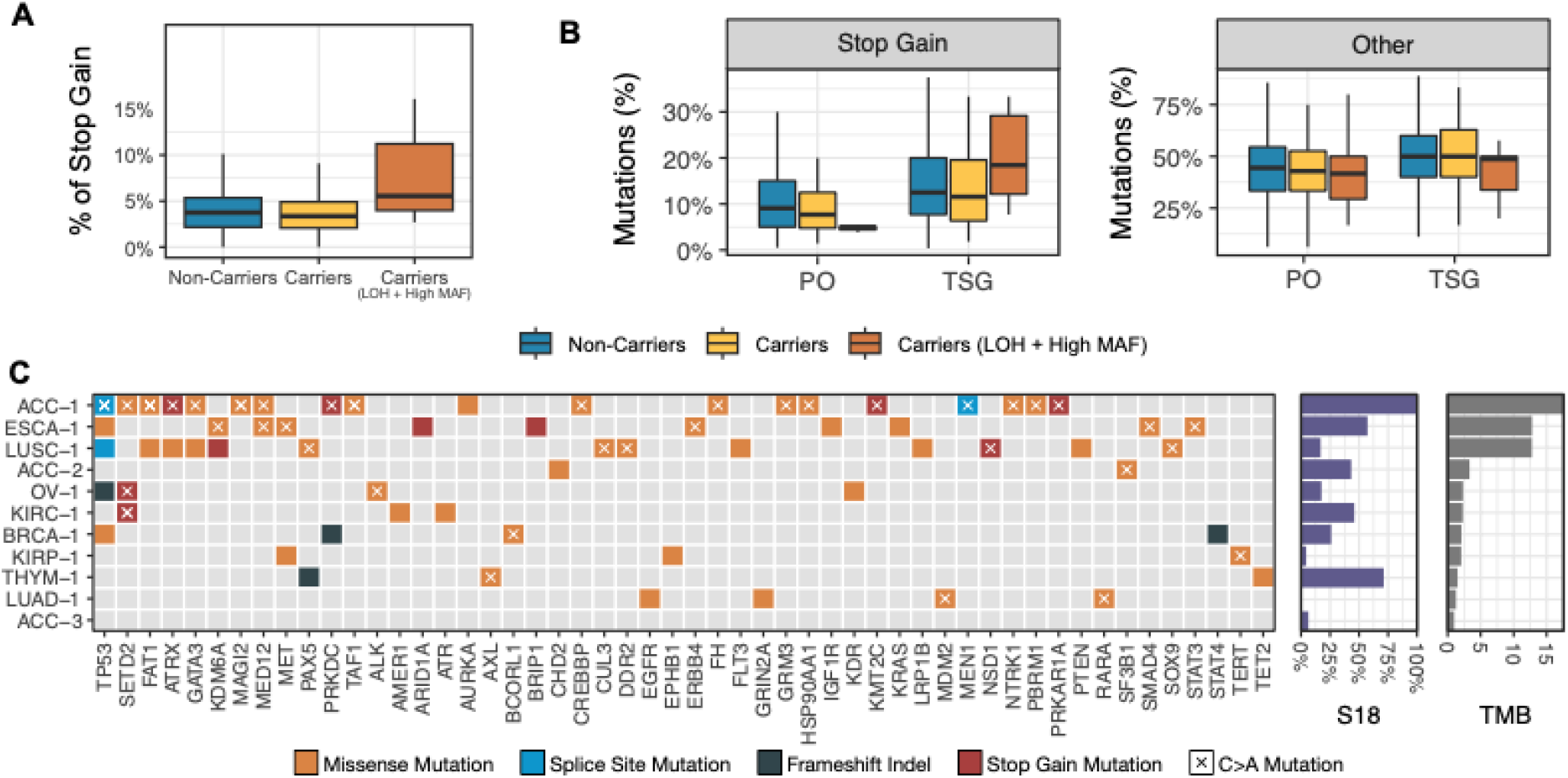
MUTYH potential stop-codon gain generation and frequent cancer gene mutations in the MUTYH variants carriers with LOH and High MAF. (A) Proportion of stop gain generating mutations in TCGA cohort. (B) Proportion of stop gain and other non synonymous mutations on cancer genes within TCGA’s patients (PO: proto-oncogene, TSG: Tumor Suppressor Gene). (C) Co-mutation plot for the 10 damaging MUTYH variant carriers with LOH for the MUTYH region and with High MUTYH MAF in the tumor’s RNA-seq quantification. Only displaying non-synonymous mutations in the cancer gene list evaluated by the FoundationONE panel. One sample (ACC-3) was removed because it lacked any cancer gene mutation. The right panel depicts COSMIC Mutational Signature 18 (S18) and Tumor Mutational Burden associated with each patient (Mutations/Mb).

To evaluate the tumorigenic potential of these stop codon gain mutations, investigated their occurrence in the COSMIC’s Cancer Gene Consensus [25]. Figure 5B shows a higher proportion of stop codon gain on tumor suppressor genes in the MUTYH deleterious variant carriers with LOH and high MAF and a decreased proportion of stop gain generation on proto-oncogenes. Thus, these results may indicate that stop gain generation is a potential tumorigenic molecular mechanism in cells carrying only a defective MUTYH copy.

### 2.6 Somatic mutation profiling of tumors from individuals carrying MUTYH damaging variants

Finally, to better understand the scope of somatic mutations in patients with *MUTYH* germline damaging variants, we investigated genes commonly used in next-generation sequencing based diagnostic tests. We analyzed somatic mutations in cancer genes (Table S5) from tumors of patients with MUTYH damaging variant, LOH for MUTYH region and high MAF on RNA-seq (Figure 5C). Only one sample (ACC-3) did not display any somatic mutation on the 400 analyzed genes. As in a real molecular test, we also evaluated the tumor mutational burden (TMB), but we found no correlation between a high TMB and a high contribution of Signature 18. Notably, we found a high number of somatic mutations in two key cancer genes: *TP53* (5 out of 11 tumors) and *SETD2* (3 of 11), which are C>A mutations and may be a consequence of a defective MUTYH.

## DISCUSSION

We prospected the largest dataset of cancer and control samples ever available to investigate the cancer risk of individuals carrying germline MUTYH defective variants. MUTYH is a base excision repair protein that prevents mutations in DNA associated with 8-oxoguanine by catalyzing the removal of adenine from inappropriately formed OG:A base-pairs. MUTYH-associated polyposis (MAP) occurs when both *MUTYH* alleles bear damaging germline variants. MAP patients present a higher risk of developing tumors, most frequently colorectal cancer [6,7]. As MAP presents itself only in an autosomal recessive fashion, far less is known about monoallelic MUTYH variant carriers and the risk of cancer development. Although some studies investigated these carriers [10,11], a more comprehensive analysis utilizing the available public tumor databases, such as the one proposed in this study, was still lacking.

Our results indicate that the frequency of deleterious MUTYH monoallelic variants carriers, although rare, is elevated in the oncologic cohort (TCGA) in comparison to the overall population, represented by the gnomAD cohort. This enrichment is not uniformly distributed across the 33 tumor types analyzed, being most prevalent on adrenocortical carcinoma, esophageal carcinoma and sarcoma.

As described for most tumor suppressor genes, the deficiency of only one allele isn’t enough to initiate a tumorigenic process [19], so it was important to carry out a systematic evaluation of both MUTYH alleles. In our query, we did not find any homozygous or compound heterozygous individuals for germline deleterious MUTYH variants, probably because the phenotype confers a specific syndrome (MAP) that would remove the individuals from the initial cohort. Also, we couldn’t find any TCGA case with both germline and somatic deleterious MUTYH allele. So, we investigate the LOH of MUTYH region as the LOH event (by aneuploidy, copy-neutral LOH or epigenetic factors) can also interrupt MUTYH functioning copy expression [26]. Our results shows that LOH is a key event to MUTYH tumorigenis contribution, once samples classified with LOH for MUTYH region and high expression of MUTYH deleterious copy displayed a much higher proportion of presence of MUTYH’s correlated signature (91%), while monoallelic carriers and non carriers had a smaller and equivalent amount of samples with the signature (23% and 25%).

We also demonstrate that cancer samples of monoallelic MUTYH variant carriers presented MUTYH related mutational signature only when accompanied by loss of the functional allele, as it was previously reported in MAP patients [27,28]. In line with studies in MAP patients [4], we outlined an association of the presence of MUTYH defect related signature with translational stop codon generation, specially on tumor suppressor genes, which may contribute to the tumorigenesis processes underlying MUTYH repair deficiency.

In our study, monoallelic MUTYH carriers presented a strong association with adrenocortical carcinoma, as it was described in other studies [23,29]. One possible factor that could help explain this observation could be the high frequency of LOH events present in this tumor type, which appears to be a key mechanism to the MUTYH tumorigenesis pathway [30]. Also, the identification of MUTYH related tumorigenis in adrenocortical carcinoma patients could be important not only to a better understating of the tumor underlying origin, but also to evaluate treatment options since some studies describe a relationship between MUTYH mutations and immunotherapy outcome [31–33].

In summary, this work performs a comprehensive investigation of the presence of germline heterozygous deleterious MUTYH among the ten thousand patients reported in TCGA. We found that, although rare, monoallelic MUTYH deficiency can trigger tumorigenesis when there is loss of the functional allele. This is an exploratory study and the findings presented here should be confirmed in further and independent research.

## METHODS

### Data availability

From oncologic patients from The Cancer Genome Atlas (TCGA), germline variants were obtained from [13], somatic mutations were obtained from Multicenter Mutation Calling in Multiple Cancers (MC3) project [34], loss of heterozygosity was obtained from Taylor et al [20] and cosmic signatures were obtained from mSignatureDB [35]. Germline allele frequencies from exome non-cancer samples from gnomAD v2.1.1 were obtained through ANNOVAR [36];

### Identification of MUTYH Germline Pathogenic Variants in TCGA and gnomAD cohort

Germline variant data from the TCGA cohort was obtained from Huang, K. et al. (2018) (10.1016/j.cell.2018.03.039. The MUTYH variants were extracted from this whole set by chromosomal region according to Gencode 19 (https://www.gencodegenes.org/). Variants were annotated using ANNOVAR [36]. Then, the set of MUTYH variants were further filtered in order to extract the variants with relevant impact on the protein structure (i.e., pathogenic/damaging variants). This multiple step filtering process consists in evaluate for each variant (1) the presence of Pathogenic/Likely Pathogenic on ClinVar [14], (2) if it was classified as damaging on functional essay [37]; (3) if it was classified as damaging/deleterious by 2 out of 3 the prediction algorithms: SIFT [15], PolyPhen2 [16] and REVEL [17]. If the variant satisfied the filtering step (1) or (2) it was classified as Tier 1 (more confident pathogenicity), variants that achieved the filtering step (3) were classified as Tier 2. After the extraction of the variants, the TCGA cohort could be classified as carrier and non-carriers. Individuals of the gnomAD v2.1.1 non-cancer cohort [18] were also classified as carriers or non-carriers for the same MUTYH variants set obtained in the TCGA cohort.

### Comparison MUTYH pathogenic variant frequency between TCGA and gnomAD cohort

In order to investigate if the proportion of MUTYH pathogenic germline carriers were different between the tumor cohort (TCGA) and the non-cancer cohort (gnomAD; v2.1.1) contingency tables carrier/non-carrier was performed to both datasets and Fisher’s exact test was performed. The calculation of the Risk Ratio was conducted considering a 95% confidence interval to evaluate multiple tumors.

### Mutant Allele frequency on RNA-seq

To understand the proportion of MUTYH transcribed mRNA molecules harbouring our specific damaging variants set we extracted the reads aligned to the MUTYH region from the TCGA carrier patients’ RNA-seq data using the provided TCGA API and slicing tools (https://docs.gdc.cancer.gov/API/Users_Guide/BAM_Slicing/). For each sample, we counted the number of mutated reads with the reference and alternative allele (corresponding to the patient’s germline variant), and calculated the MAF as: MAF = N_ALT alleles_ / Coverage.

### Mutational Signature Analysis

For each sample of TCGA cohort all signatures from Cosmic Mutational Signatures v2 [38] were obtained from mSignatureDB [35]. Next, the mean contribution from each signature between MUTYH pathogenic germline TCGA carriers and non-carriers were compared using the Mann–Whitney U test.

### Loss of heterozygosity

Loss of heterozygosity (LOH) was obtained from ABSOLUTE analysis on TCGA’s samples available in Taylor et al. [20]. We considered LOH samples the ones with LOH region that covered the entire MUTYH sequence (chr1:45794835-45806142) on the provided data.

### Tumor Mutational Burden

Tumor Mutational Burden (TMB) was calculated as the number of all mutations (including synonymous mutations) present in the TCGA somatic mutation data set from MC3 (10.1016/j.cels.2018.03.002) divided by the estimate covered exome region (38Mb).

### Statistical Analysis & Figures

All statistical analysis was performed in R (v.4.0.3). Figures were generated using ggplot2 R package with some manual aesthetic adjustments when necessary.

## Data Availability

We have used only public data.

## Acknowledgment

The authors acknowledge the National Council for Scientific and Technological Development (CNPq) and the São Paulo Research Foundation (FAPESP, grants 2018/15579-8, 2019/04927-8 and 2020/06091-1) for financial support.

## Conflicts of Interest

The authors declare no conflict of interest

## Notes

### Competing Interest Statement

The authors have declared no competing interest.

### Funding Statement

The authors acknowledge the National Council for Scientific and Technological Development (CNPq) and the Sao Paulo Research Foundation (FAPESP, grants 2018/15579-8, 2019/04927-8 and 2020/06091-1) for financial support.

